# Publication trend, impact and performance in mental health during and post COVID-19 pandemic

**DOI:** 10.1101/2024.07.04.24309983

**Authors:** Nurhidayah Bahar, Fadilla ‘Atyka Nor Rashid, Nor Diana Ahmad

## Abstract

This study presents a bibliometric analysis of publications on mental health during and post-COVID-19 pandemic in Malaysia. The dataset comprises 167 documents retrieved from the Scopus database covering the period from 2020 to 2022. Using the open-source tool Bibliometrix for science mapping analysis, we examined various aspects of these publications, including document type, number of articles, total citations, most relevant sources, source impact, most relevant authors, affiliations, globally cited documents, and a word cloud. Our findings reveal a significant and continuous growth in mental health-related publications since the onset of the pandemic in 2020, highlighting an intensified focus on this critical area. This surge in research emphasizes the heightened importance of understanding and addressing mental health issues exacerbated by the pandemic. By providing a comprehensive summary of the bibliometric data, this study enhances our understanding of publication trends and the evolving landscape of mental health research, offering valuable insights for researchers, policymakers, and practitioners aiming to respond to mental health challenges in the post-pandemic era.

## Introduction

Mental health can be regarded as a state of mental well-being that enables people to cope with the stresses of life, realize their abilities, learn well and work well, as well as contribute to their community [1]. Therefore, mental health is one of many fundamental human rights and it is important for the development of personal well-being, community and socio-economic. Mental health conditions include mental disorders, psychosocial disabilities and other mental states. Some of determinants that can make people more vulnerable to mental health problems are individual psychological and biological factors (i.e., emotional skills), substance use and genetics, in a state of unfavourable economic, social, and environmental circumstances (e.g., poverty, violence, and inequality) [1].

Mental disorder also referred to as a mental illness or psychiatric disorder refers to disturbance in an individual’s thinking, emotional regulation, or behaviour. According to a report published by Global Health Data Exchange (GHDx) in 2019, 970 million people around the world were living with a mental disorder, that is equivalent to 1 in every 8 people [2]. The number of people living with any sort of metal disorder rose significantly during COVID-19 pandemic in 2020. World Health Organization (WHO) in one of its article estimated 26% and 28% increase respectively for anxiety and major depressive disorders in just one year [3,4]. WHO also listed different types of mental disorders including anxiety disorders, depression, bipolar disorder, post-traumatic stress disorder (PTSD), schizophrenia, eating disorders, neurodevelopmental disorders, disruptive behaviour and dissocial disorders [3].

The literature review shows that there is a significant interest in investigating the implications of COVID-19 towards mental health. A closer look to the literature on mental health during and post COVID-19 pandemic in the Malaysian context, reveals a great number of studies have reported groups of people that psychological affected due to the global pandemic since its outbreak in late December 2019. Most of the earlier studies research the impact towards the well-being of healthcare workers [5–7] and their family members [8]. Healthcare workers play a vital role in management of the COVID-19 pandemic and often working longer hours to combat the outbreak. This caused a heavy-toll on their family members especially their children.

Psychological impact of COVID-19 were also observed among students at different levels such as primary and secondary school [9,10] as well as university students [11,12]. Apart from student population, researchers also extend the investigation to teachers [13], patients [14], parents [15] and elderly [16]. Motivated by this trend, present study aims to explore the current state of publication in mental health during and post COVID-19 pandemic. Particularly, the objectives of this study include (i) to examine the productivity of research through the publication trends and (ii) to evaluate its impact and performance using a few metrics such as the total citations, citations per paper, citations per year, number of cited papers, h-index, g-index and m-index.

## Methods

This study employed bibliometric analysis to explore publication trend, impact and performance of studies relating to mental health during and post COVID-19 pandemic in Malaysia. Fig 1 shows the steps of how the dataset was obtained from the Scopus database on October 13, 2022.

**Fig 1.**
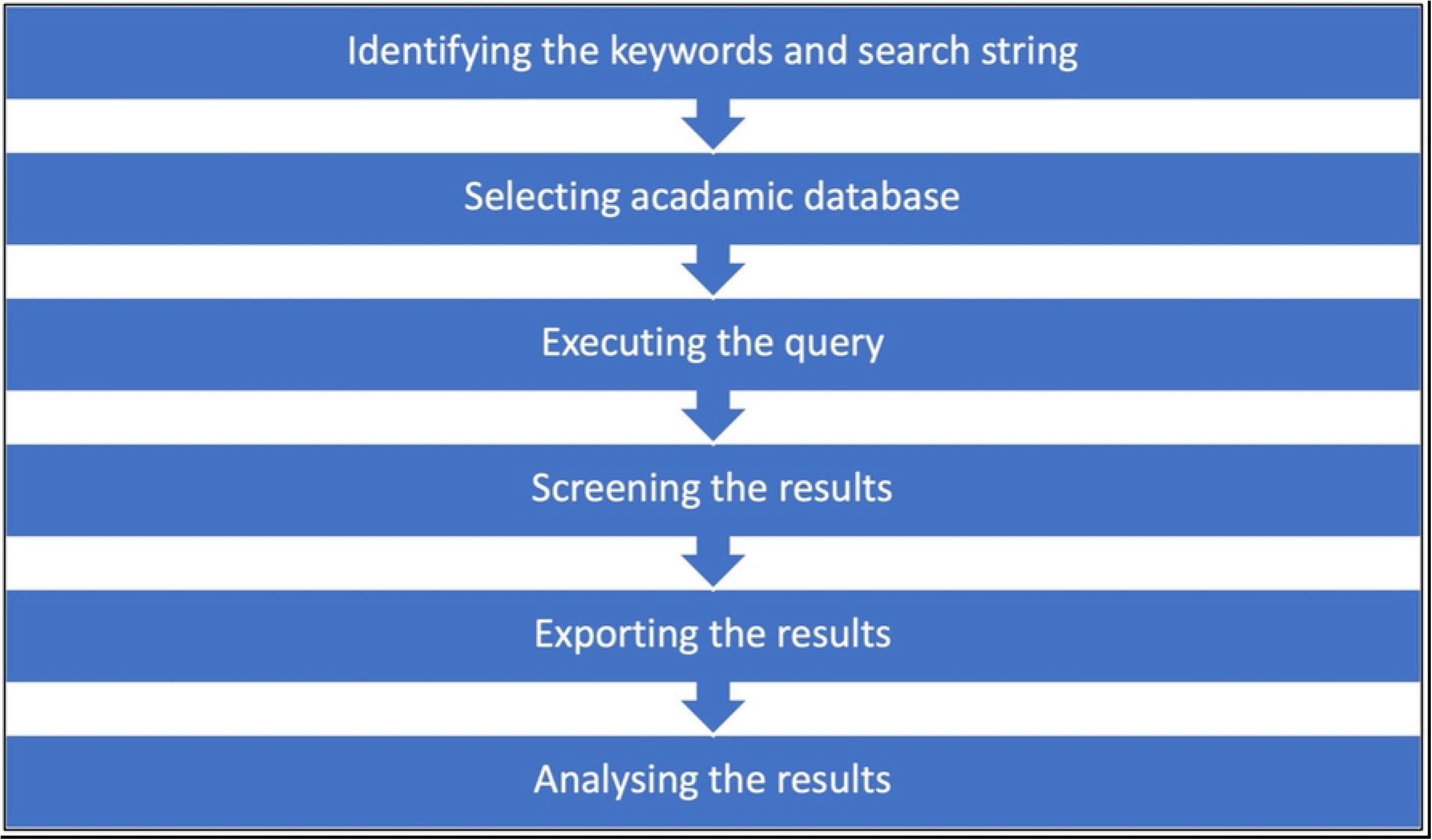
Steps undertaken to perform bibliometric analysis

First, the keywords including other words that shared similar meanings to the topic were identified. For example, ‘mental health’, ‘covid’ and ‘pandemic’. Second, the academic database that has bibliographic information such as document title, authors, year, source, publication stage, and citation was chosen. This study chose to analyse bibliographic information from Scopus because it is one of the largest searchable citation and abstract literature search list [17].

Once the academic database was selected, the query was executed using suitable search strategy for Scopus. Fig 2 shows the query used to search for relevant articles that is significant with the research area and the aim of the study. The query were searched within article title, abstracts and keywords revealed 167 document results. Next, screening was performed to ensure all of the documents are suitable to be included in the analysis. None of the document was removed and the final number of documents to be analysed remained 167.

**Fig 2.**
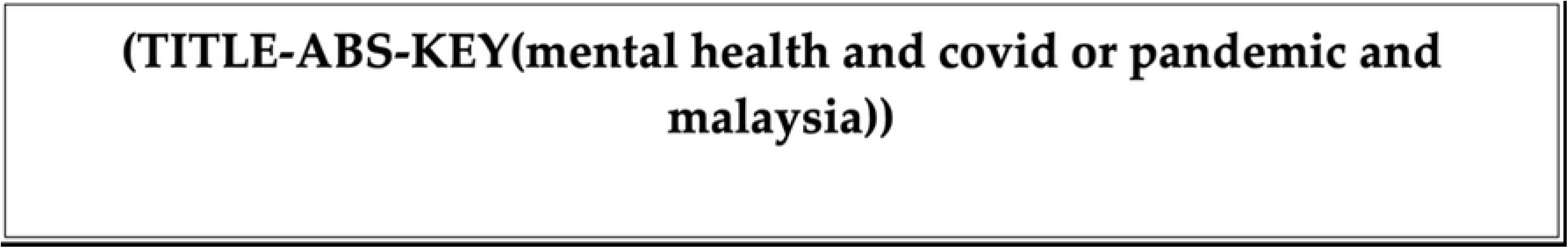
The query

The documents were published from 2020 to 2022 and derived from different Open Access types available in Scopus including Gold, Hybrid Gold, Bronze and Green. There were 160 documents that can be categorized as published article and another seven documents are article in press. This study used Bibliometrix, an R-tool to comprehensively perform the mapping analysis. Particularly, Biblioshiny was used to generate the visual presentation (e.g., list, graphs, tables, and word cloud) of the gathered data. This tool was widely used in many bibliometric analysis studies [18].

## Results

A total of 167 documents were identified from the Scopus database published in three years; 2020 (61 documents), 2021 (82 documents), and 2022 (24 documents). There were 13 types of documents published relevant to this topic including article (143 documents), review (9 documents), conference paper (7 documents), letter (5 documents), editorial (2 documents), and book chapter (1 document). All of these documents were published in 98 source titles including journals, conference proceeding, and book series involving 881 authors. A total of 5 documents were single-authored publications while the remaining documents were multi-authored publications. Thus, Therefore, the degree of research collaboration for this subject is high (97%). The average co-authors per document is 6.45 and average citations per doc within the timespan (2020:2022) is 7.054. The percentage of international co-authorship is 38.32 percent. Fig 3 shows the three field plot consists of three parameters namely authors’ country at the left field, authors at the middle field and authors’ affiliation at the right field. This three field plot, however, depicts only twenty records for each parameter. This diagram helps to visualize the relationship between these three parameters.

**Fig 3.**
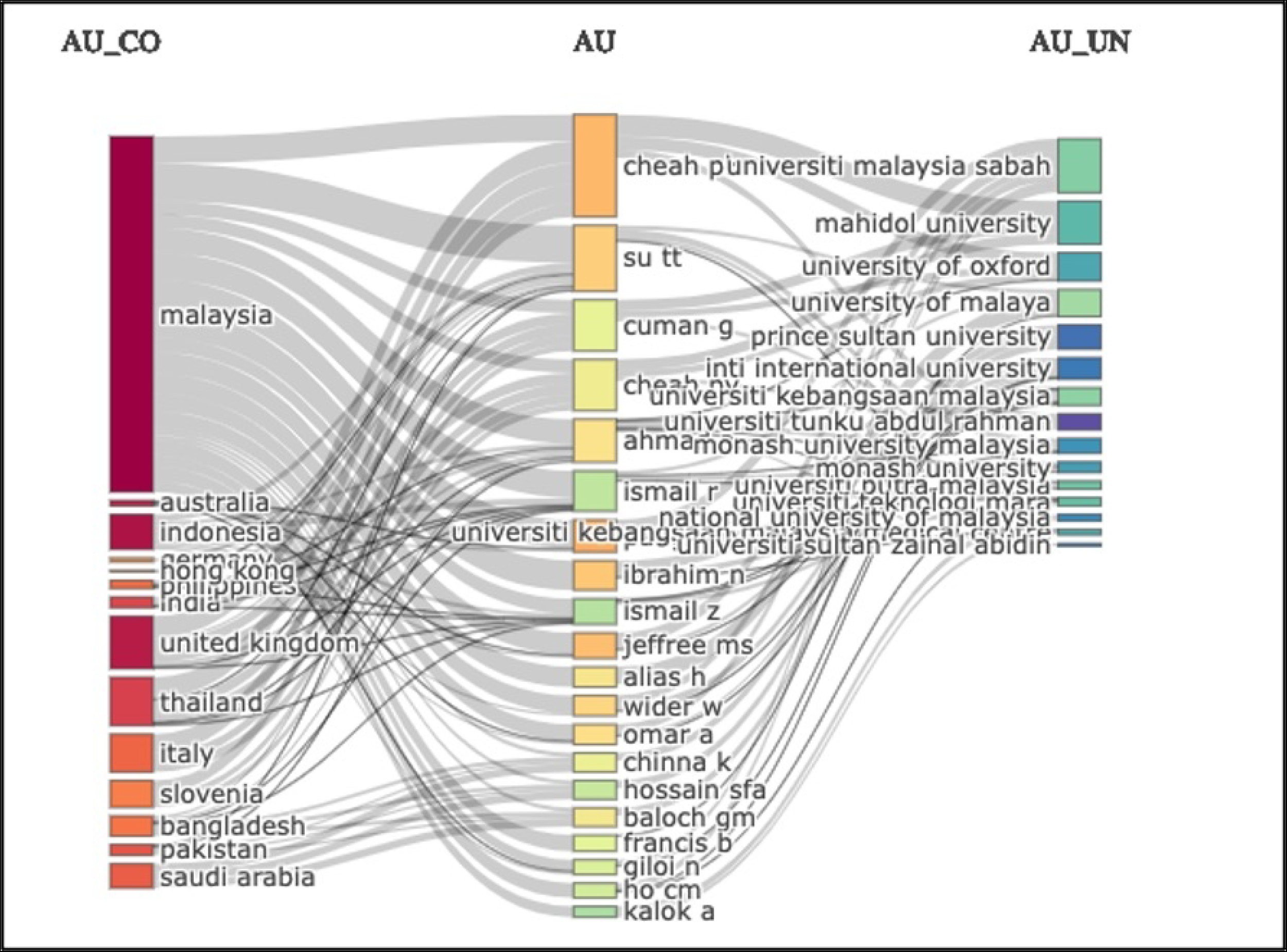
Three field plot

### Growth of publications

Growth of the publications was examined based on number of articles and total citations. Table 1 summarises the details statistic of annual publications on the dataset including number of articles, mean total citations per article, mean total citations per year and citable years. There is a growth of publication since 2020 and year 2022 with 59.43% of annual growth rate. Year 2020 has the highest average value for both categories namely mean total citation per article (23.58) and mean total citation per year (11.79).

**Table 1.**
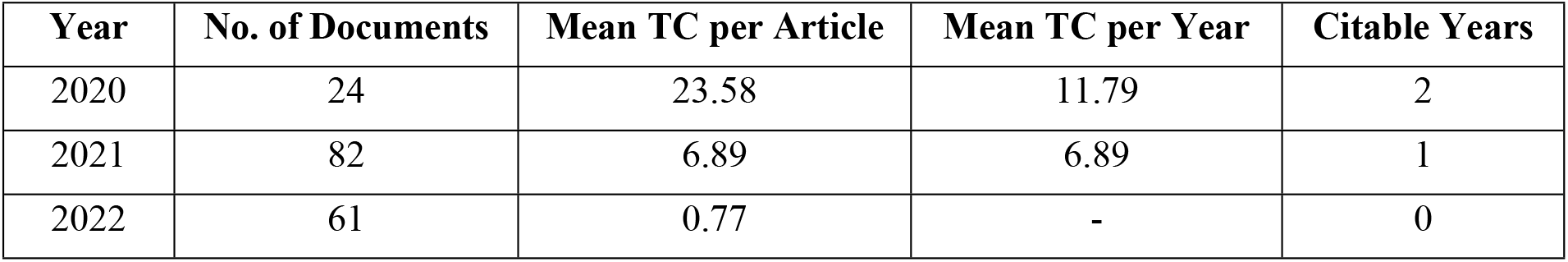
Number of articles and total citations (TC)

### Relevant Sources and Source Impact

The main three sources for this dataset are journal, review and conference. Table 2 present the title of top 10 relevant sources that have published the documents related to mental health during and post COVID-19 pandemic from 2020 to 2022. These source titles were ranked based on the number of publications related to the search area. This analysis shows the number of articles being published, performance and impact that were measured based on four metrics including total publications (TP), total citations (TC), h-index, g-index, and m-index. International Journal of Environmental Research and Public Health has the highest number of publications (21 documents) and highest total citations (398). On the other hand, Frontiers in Psychology has the least number of publications (3 documents) and lowest total citations.

**Table 2.** Top 10 relevant sources.

| Sources | TP | TC | h-index | g-index | m-index |
| --- | --- | --- | --- | --- | --- |
| International Journal of Environmental Research and Public Health | 21 | 398 | 8 | 19 | 2.667 |
| Plos One | 11 | 237 | 5 | 11 | 2.500 |
| Frontiers in Public Health | 5 | 1 | 1 | 1 | 0.500 |
| Malaysian Journal of Public Health Medicine | 5 | 6 | 1 | 2 | 0.500 |
| Medical Journal of Malaysia | 5 | 10 | 2 | 3 | 0.333 |
| Sustainability | 4 | 21 | 2 | 4 | 1.000 |
| Frontiers in Psychiatry | 3 | 30 | 1 | 3 | 0.500 |
| Frontiers in Psychology | 3 | 2 | 2 | 2 | 1.000 |
| Malaysian Journal of Medicine and Health Sciences | 3 | 5 | 1 | 2 | 0.500 |
| The Lancet Psychiatry | 3 | 7 | 2 | 2 | 1.000 |

### Most Relevant Authors

Most relevant authors was analysed based on the number of articles produced by them and articles fractionalised as shown in Table 3. This analysis quantifies the productivity of top 10 authors derived from this dataset. Nicholas Pang Tze Ping from Universiti Malaysia Sabah (UMS) has the highest number of publications and positioned at the top of the list, followed by Cheah Phee Kheng and Mohammad Saffree Jeffree with 6 articles. The results also show collaboration opportunities for other researchers who wish to pursue studies in this area.

**Table 3.**
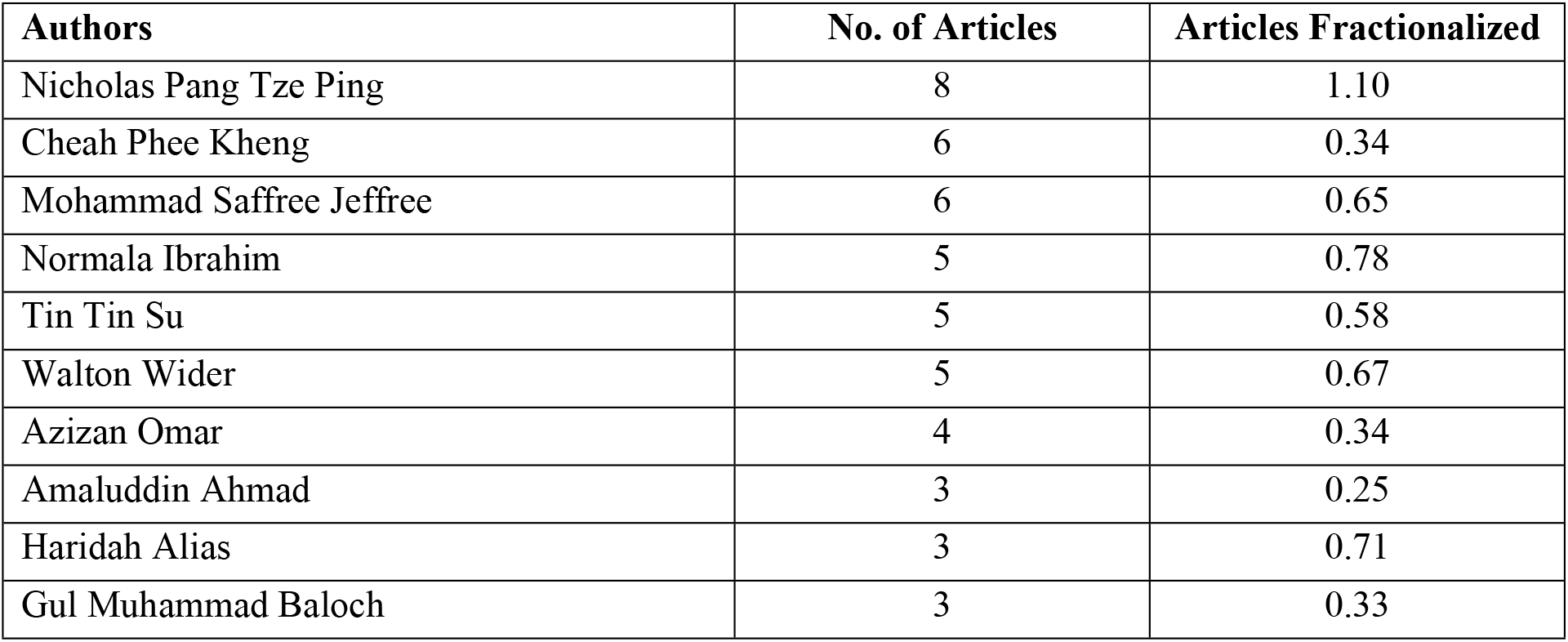
Top 10 most relevant authors.

### Most Relevant Affiliations

The dataset was further analysed by examining the frequency of publications according to most relevant affiliations. Most relevant affiliations refer to the productivity of publications based on the institutions where the authors claimed in the publication [19]. Table 4 depicts the top 10 institutions that has contributed in publishing documents related to mental health studies during and post COVID-19 pandemic. Universiti Sains Malaysia is at the top of the list (34 articles), followed by Universiti Kebangsaan Malaysia and Universiti Malaya (26 articles). The rest of institutions have published less than 20 articles from 2020 to 2022 such as Universiti Teknologi MARA, Universiti Malaysia Sabah, Universiti Putra Malaysia to name but a few.

**Table 4.**
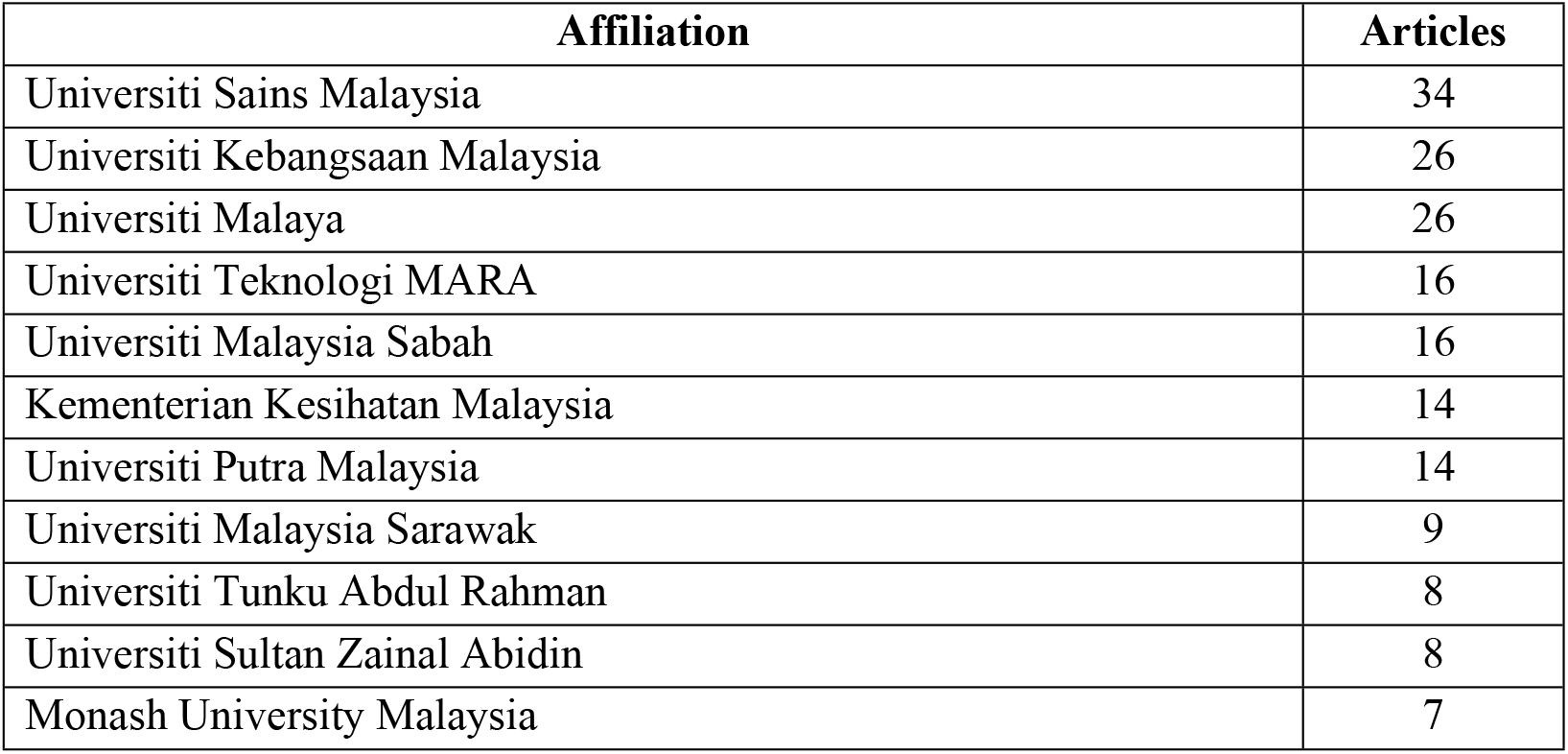
Top 10 most relevant affiliations.

| Affiliation | Articles |
| --- | --- |
| Universiti Sains Malaysia | 34 |
| Universiti Kebangsaan Malaysia | 26 |
| Universiti Malaya | 26 |
| Universiti Teknologi MARA | 16 |
| Universiti Malaysia Sabah | 16 |
| Kementerian Kesihatan Malaysia | 14 |
| Universiti Putra Malaysia | 14 |
| Universiti Malaysia Sarawak | 9 |
| Universiti Tunku Abdul Rahman | 8 |
| Universiti Sultan Zainal Abidin | 8 |
| Monash University Malaysia | 7 |

### Most Global Cited Documents

Most global cited documents was calculated based on the number of citations received by the particular document as of the search was undertaken from all the documents around the world [19]. The citations were count based on all articles within the Scopus database that cite the paper. These most global cited documents were measured by total citations (TC), total citations per year and normalized TC (NTC) as tabulated in Table 5. Document by Sheela Sundarasen et al. [20] entitled “Psychological Impact of COVID-19 and Lockdown among University Students in Malaysia: Implications and Policy Recommendations” received the highest number of citations, followed by article entitled “The impact of COVID-19 pandemic on physical and mental health of Asians: A study of seven middle-income countries in Asia” written by Cuiyan Wang et al. in 2021.

**Table 5.**
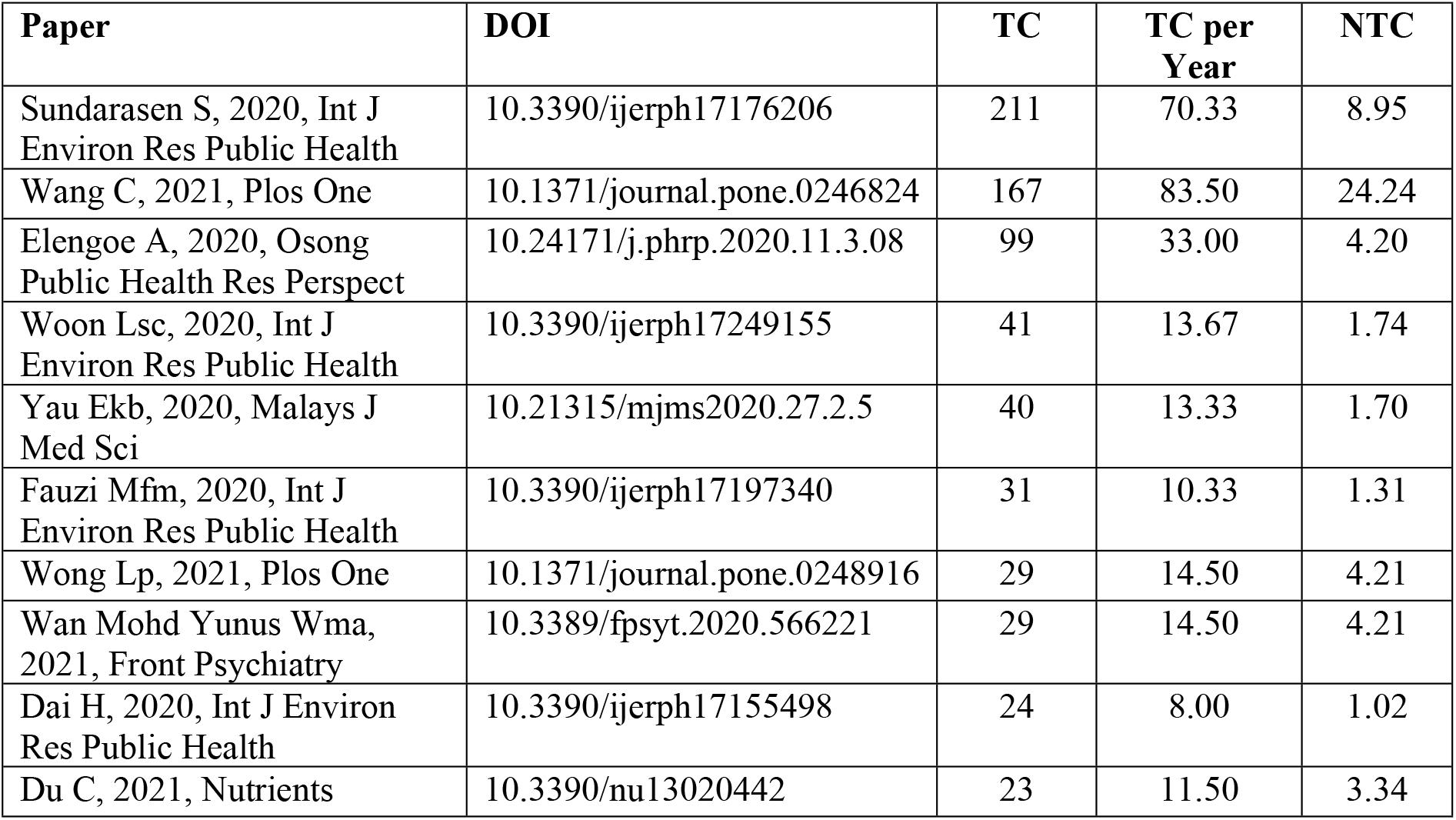
Most global cited documents.

### Keyword and Word Cloud

There are several important keywords used in this dataset. The highest number of keyword used is COVID-19 or some documents might also use Coronavirus Disease 2019. Some other keywords that frequently used in the dataset are human/humans, pandemic, mental health, female and adult. This list of keyword was directly retrieved from Scopus. Alternatively, word cloud was generated to examine word frequency in the entire dataset. Fig 4 is a graphical representation of word frequency that emphasises words that frequently appeared in the dataset. This visual representation gives greater rank to words that appear more frequently. From this figure it can be seen there are a few words were highlighted such as depression (95 occurrences), pandemic (94 occurrences), female (93 occurrences), male (85 occurrences), and anxiety (74 occurrences). This is consistent with the keywords listed by Scopus.

**Fig 4.**
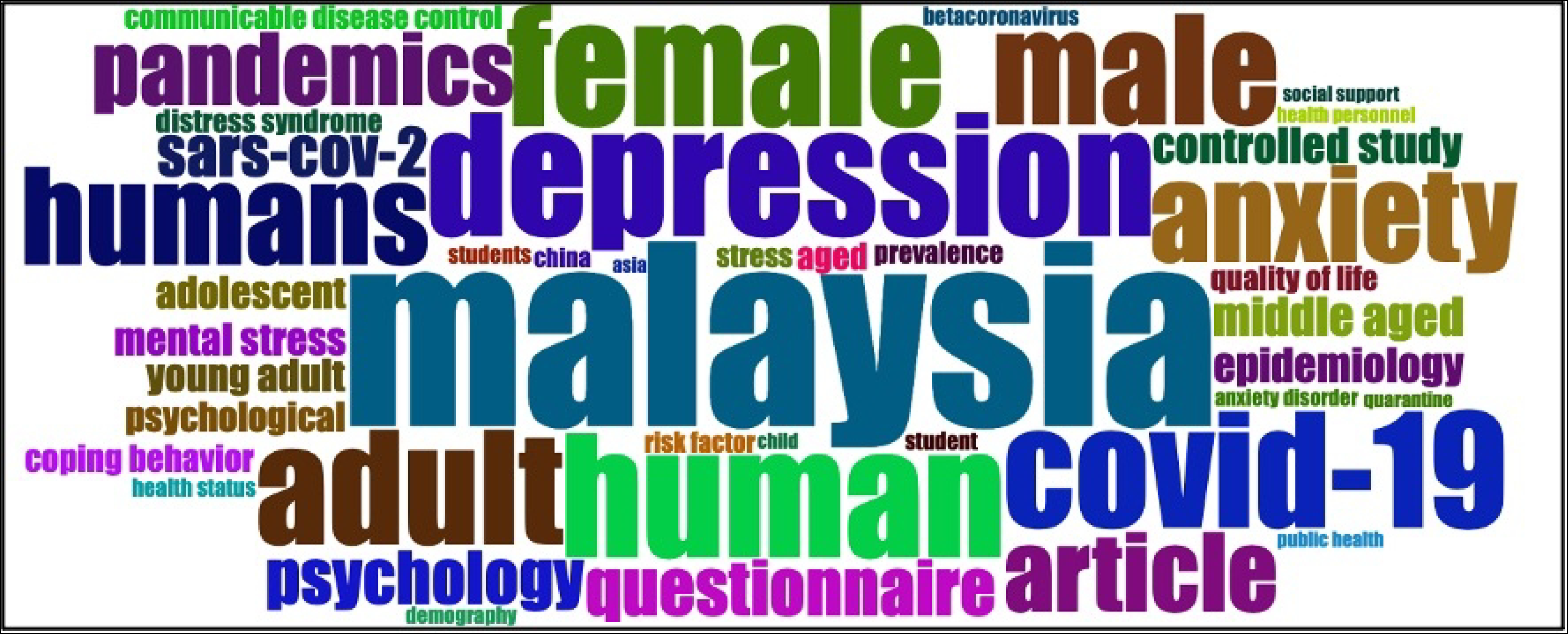
Word cloud of authors’ keywords

## Discussion

The first cases of COVID-19 in Malaysia were reported in late January 2020. In March 2020, the Malaysian government imposed the Movement Control Order (MCO), which included restrictions on movement, and the closure of non-essential businesses and schools. Subsequently, MCOs were implemented in various forms (e.g., Conditional MCO, Enhanced MCO) based on the severity of outbreaks. Public health interventions such as mask mandates, social distancing, travel restrictions, and testing protocols were enforced throughout the period. The COVID-19 pandemic had a profound impact on mental health in Malaysia. Factors contributing to mental health challenges included social isolation and loneliness due to lockdowns and movement restrictions [21], uncertainty and anxiety about the virus and health [22], economic stress from job losses and reduced income as well as grief and trauma from illness and deaths within communities [23].

Given the significant mental health challenges faced during this period, it is crucial to understand the corresponding research trends. To answer the research question (What is the current state of publication in mental health during and post COVID-19 pandemic?), this study examined the publication trend by analysing the bibliographic data collected including document type, number of articles and total citations, most relevant sources, source impact, most relevant authors, most relevant affiliation, most global cited documents, and the word cloud. The selection of articles between the year 2020 until 2022 in this bibliometric analysis is significant to further understand the publication trend, impact and performance in mental health during and post COVID-19 pandemic. The findings suggest that there is a significant growth of publication for studies relating to mental health especially during and post pandemic.

The studies involved individuals with diverse profiling such as students[12,24], teachers[25], parents[15], and healthcare workers[26]. It is evident that the findings from these studies are deemed important highlighting well-being mental health among Malaysian during pandemic period. Most of the studies were conducted during the early stage of COVID-19 pandemic in Malaysia, thus, the findings reported in this studies shed light on the issues involved within this area. For instance, researchers developed a tool to measure mental health conditions among Malaysians [24], determined the distress symptoms and the coping strategies among medical students [12] as well as investigated the prevalence and severity of depression, anxiety, and stress healthcare workers during the covid-19 pandemic [5].

The rise in the number of publications on mental health in 2021 can be attributed to several factors directly related to the evolving COVID-19 situation. As the pandemic progressed into its second year, the cumulative effects of prolonged social isolation, economic uncertainty, and health-related anxiety became more pronounced. Researchers and healthcare professionals increasingly recognized the urgent need to understand and address the mental health crisis derived from these conditions. The acceleration of vaccination programs in early 2021 also played a role. As vaccines became widely available and public health measures evolved, researchers began to focus on the mental health implications of vaccination, vaccine hesitancy, and the transition to a new normal[27–30].

## Conclusion

This study aims to extract meaningful information about the structures of publication trend in mental health during and post-pandemic from the Scopus database. It is clear that there is positive growth of scientific publications in this topic since 2020. The findings from this study provide several contributions. First, the output from the bibliometric analysis could help to evaluate the journals that are desirable for the research committee to publish/subscribe. Second, the results provide insights relating to trend, performance and the state of the art of publication within the field of present study. Third, this study would benefit policymaker or funding sponsor for deciding allocation of the fund, developing research guidelines and interest, and strategizing the national agenda based on the report that shows ranking of education institutions and the profiling of the researchers’ productivities and impact. More importantly, the present study help other researchers to understand the global issue regarding mental health and propose pathways for further studies. The growing mental health problem has significantly affected people worldwide, including in Malaysia. Thus, there is an urgent need for effective interventions and robust policies, involving key players such as healthcare providers, policymakers, and community organizations, to manage mental health disorders among the population.

There are a few limitations to this bibliometric study. First, the data presented in this study are derived solely from the Scopus database and were collected using specific keywords. While Scopus provides a comprehensive source of academic literature, reliance on a single database and the use of specific keywords may have constrained the breadth and depth of the analysis. Alternative databases and additional or broader keywords could potentially yield different results and perspectives, thereby enhancing the comprehensiveness of future studies on this topic. Second, the analysis is limited to bibliographical information. Further analysis can be conducted to examine the collaborations networks within the reference clusters and relationships, scientific communities, research themes and evolution of keywords or terms, and research gaps. Future research in this field could explore several promising directions. Firstly, investigating collaboration networks among researchers in mental health during and post-COVID-19 could provide insights into interdisciplinary approaches and effective knowledge dissemination strategies. Additionally, identifying research gaps within specific subfields of mental health, such as interventions for vulnerable populations or the impact of socioeconomic factors, could guide targeted research efforts and policy initiatives. Furthermore, analyzing the evolution of research themes over time, particularly in relation to emerging global health crises or shifts in public health policies, could offer valuable perspectives on the dynamic nature of mental health research. These future directions not only expand upon the current findings but also contribute to enhancing our understanding and addressing the complex challenges posed by mental health in a post-pandemic world.

## Data Availability

All relevant data are within the manuscript and its Supporting Information files.

NA

## Acknowledgements

Do not include funding or competing interests information in Acknowledgments.

